# SARS-CoV-2 and the Role of Orofecal Transmission: Systematic Review

**DOI:** 10.1101/2020.08.04.20168054

**Authors:** C Heneghan, EA Spencer, J Brassey, T Jefferson

## Abstract

**Background:** How SARS-CoV-2 is transmitted is of key public health importance. SARS-CoV-2 has been detected in the feces of some Covid-19 patients which suggests the possibility that the virus could additionally be transmitted via the orofecal route.

**Methods:** This review is part of an Open Evidence Review on Transmission Dynamics of Covid-19. We conduct ongoing searches using LitCovid, medRxiv, Google Scholar and Google for Covid-19; assess study quality based on five criteria and report important findings on an ongoing basis. Where necessary authors are contacted for further details or clarification on the content of their articles.

**Results:** We found 59 studies: nine reviews and 51 primary studies or reports (one cohort study also included a review) examining the potential role of orofecal transmission of SARS-CoV-2. Half (n=29) were done in China. Thirty seven studies reported positive fecal samples for SARS-CoV-2 based on RT-PCR results (n=1,034 patients). Six studies reported isolating the virus from fecal samples of nine patients, one study isolated the virus from rectal tissue and one laboratory study found that SARS-CoV-2 productively infected human small intestinal organoids. Eleven studies report on fecal samples found in sewage, and two sampled bathrooms and toilets.

**Conclusions:** Various observational and mechanistic evidence support the hypothesis that SARS-CoV-2 can infect and be shed from the human gastrointestinal tract. Policy should emphasize the importance of strict personal hygiene measures, and chlorine-based disinfection of surfaces in locations where there is presumed or known SARS-CoV-2 activity.

## Background

Understanding how, when and in what types of settings SARS-CoV-2 spreads between people is critical to developing effective public health and infection prevention measures to break chains of transmission.^1^ Current evidence suggests SARS-CoV-2 is primarily transmitted via respiratory droplets and aerosols; and can occur between pre-symptomatic or symptomatic infected individuals to others in close contact.

SARS-CoV-2 has been shown to contaminate and survive on certain surfaces, but currently, no reports have directly demonstrated fomite to human transmission. SARS-CoV-2 has also been detected in the feces of some patients which taken together with fomite transmission suggest the possibility that SARS-CoV-2 could transmit via the orofecal route. “Orofecal” describes a route of transmission where the virus in fecal particles can pass from one person to the mouth of another. Main causes include lack of adequate sanitation and poor hygiene practices. Fecal contamination of food is another form of orofecal transmission.

## Methods

We are undertaking an open evidence review investigating factors and circumstances that impact on the transmission of SARS-CoV-2, based on our published protocol. In brief, this review aims to identify and evaluate relevant articles (peer-reviewed or awaiting peer review) that examine the mode of viral transmission and ecological variables influencing the mode of transmission. We conduct an ongoing search using LitCovid, medRxiv, Google Scholar and Google for Covid-19 for keywords and associated synonyms. Results are reviewed for relevance and for articles that looked particularly relevant forward citation matching was undertaken and relevant results were identified. Studies with modelling are only included if they report transmission outcome data and not predicted outcomes. One reviewer extracts data from the included studies that is then checked by a second reviewer and a summary is uploaded to an accessible Evidence Explorer at https://www.cebm.net/evidence-synthesis/transmission-dynamics-of-covid-19/

From included studies we extract information on the study characteristics, the population, the funding and the main methods, reported mode of transmissions and associated outcomes, We assess study quality based on five criteria and report important findings on an ongoing basis. Where necessary we write to authors of included studies for further details or clarification on the content of their articles.

## Results

We found 59 studies: nine reviews and 51 primary studies or reports (one study undertook a cohort study and included a review) examining the potential role of orofecal transmission of SARS-CoV-2 (see Table 1 of references ^w1-w59^). Half (n=29) were done in China

Table 1. References:

Amongst the 51 primary studies or reports (see Table 2. [Study Characteristics](https://docs.google.com/document/d/1bhwkX_eXxLUXmictta5RxSRUjQgCZglTjaZZ2kHbMC4/edit#)) thirty seven studies reported positive fecal samples for SARS-CoV-2 based on RT-PCR results (n=1,034 patients). Six ([Jeong](https://www.cebm.net/study/viable-sars-cov-2-in-saliva-urine-and-stool-from-covid-19-patients/), [Wang W](https://www.cebm.net/study/detection-of-sars-cov-2-in-different-types-of-clinical-specimens/), [Xiao F](https://www.cebm.net/study/covid-19-evidence-for-gastrointestinal-infection-by-sars-cov-2-2/), [Tang M](https://www.cebm.net/study/covid-19-evidence-for-gastrointestinal-infection-by-sars-cov-2-2/), [Xiao F, Sun J](https://www.cebm.net/study/infectious-sars-cov-2-in-feces-of-patient-with-severe-covid-19/), [Yoa H](https://www.cebm.net/study/impact-pathogenicity-of-sars-cov-2/) and [Zhang Y 2020](https://www.cebm.net/study/isolation-of-sars-cov-2-from-a-stool-specimen-of-confirmed-case-of-covid-19/)) report isolating the virus. Eleven studies report on fecal samples in sewage. (see [Study Characteristics](https://docs.google.com/document/d/1bhwkX_eXxLUXmictta5RxSRUjQgCZglTjaZZ2kHbMC4/edit#)) Three studies were done in laboratories ([Lamers](https://www.cebm.net/study/sars-cov-2-productively-infects-human-gut-enterocytes/), [Zang](https://www.cebm.net/study/covid-19-tmprss2-and-tmprss4-promote-sars-cov-2-infection-of-human-small-intestinal-enterocytes/) and [Zhou J](https://www.cebm.net/study/covid-19-infection-of-bat-and-human-intestinal-organoids-by-sars-cov-2/)), and two sampled bathrooms and toilets for viruses ([Ding Z](https://www.cebm.net/study/covid-19-toilets-dominate-environmental-detection-of-sars-cov-2-in-a-hospital/), [Ong](https://www.cebm.net/study/air-surface-environmental-and-personal-protective-equipment-contamination-by-sars-cov-2-from-a-symptomatic-patient/)).

Overall the quality of the evidence was low to moderate mainly due to a lack of standardisation of techniques, omissions in reporting and a failure to account for biases in the research process. (see Table 3 Quality of Included Studies).

Table 2. Study Characteristics:

Table 3. Quality of Included Studies:

## SARS-COV-2 excretion and detection via the orofecal route - evidence from reviews

The main findings of each study are summarised in Table 4.

Table 4. Table of Findings of Included Studies:

We found nine reviews with overlapping studies that assessed the excretion and detection of SARS-COV-2: six in mixed adults and children [[Parasa](https://www.cebm.net/study/prevalence-of-gastrointestinal-symptoms-and-fecal-viral-shedding-in-patients-with-covid-2019-a-systematic-review-and-meta-analysis/), ^w31^ [Amirian](https://www.cebm.net/study/potential-faecal-transmission-of-sars-cov-2/), ^W2^ [Cheung, W](https://www.cebm.net/study/sars-cov-2-infection-in-fecal-samples-from-hong-kong/)^10^ [Ding S](https://www.cebm.net/study/covid-19-is-sars-cov-2-also-an-enteric-pathogen-with-potential-fecal-oral-transmission/), ^W12^ [Gupta](https://www.cebm.net/study/covid-19-persistent-viral-shedding-of-sars-cov-2-in-faeces/) ^W15^ and [Tian](https://www.cebm.net/study/covid-19-gastrointestinal-features-in-covid-19-and-the-possibility-of-faecal-transmission/) ^W39^] - two exclusively in children [[Santos](https://www.cebm.net/study/covid-19-fecal-shedding-in-children/). ^W34^ [Donà](https://www.cebm.net/study/covid-19-fecal-oral-transmission-of-sars-cov-2-in-children/) ^W14^]. We found one review on food handling [[Kingsbury](https://www.cebm.net/study/covid-19-potential-for-foodborne-transmission-of-covid-19/) ^W20^] and [Cahill](https://www.cebm.net/study/covid-19-in-wastewater/) ^W3^ et al. did a short review on the available evidence of orofecal transmission and its possible relationship to wastewater management.

A systematic review and meta-analysis of 29 relevant studies (23 published and 6 preprints conducted between November 1, 2019, and March 30, 2020 assessed as moderate quality) including 4,805 patients. ^w31^ The review reported that approximately 12% of patients with SARS-CoV-2 had gastrointestinal symptoms, including diarrhoea, nausea, and vomiting. Eight studies reported the detection of viral RNA of SARS-CoV-2 in stool of patients confirmed by nasopharyngeal swab testing or respiratory secretion analysis for PCR to have Covid-19: pooled analysis reported RNA shedding in stool was 40.5% (95% CI: 27.4% to 55.1%; I^2^ 83%) of Covid-19 patients. A second review of fourteen small studies (n = 476; range 1 to 153 cases) reported that SARS-CoV-2 may be transmitted orofecally. ^W2^ The review does not have a methods chapter and there is only one table describing the included studies. A third review of observational and mechanistic evidence provides further support that SARS-CoV-2 can infect and be shed from the human gastrointestinal tract. ^W12^

A fourth review with 26 articles included in the final analysis with 824 patients included across the studies and 540 tested for fecal viral RNA: 291 (54%) had positive fecal RT-PCR tests. ^W15^ Of the 199 patients who tested positive for fecal viral RNA and who were followed up with stool testing, 125 (63%) had persistent shedding of virus in the stool samples after a negative nasopharyngeal swab. The duration of fecal shedding of viral RNA after clearance of respiratory samples ranged from 1 to 33 days and in one patient up to 47 days from symptom onset. Only one study tested the viability of fecally excreted viruses. Of 153 stool specimens tested in this study, four were tested for viability and two (50%) were viable.

Tian et al ^W39^ reviewed 15 included studies with data from 2,023 patients (mainly from China) and showed that gastrointestinal symptoms are common in Covid-19 patients and were reported with increased prevalence as the epidemic progressed in China. Among 2,033 Covid-19 patients, gastrointestinal symptoms were observed in 3% (1/41) to 79% (159/201) of the cases. Symptoms included anorexia 40%; diarrhoea 2%; vomiting 3.6%; nausea 1%; abdominal pain 2.2%; and gastrointestinal bleeding 4%.

Diarrhoea was the most common gastrointestinal symptom in children and adults, with a mean duration of 4.1 ± 2.5 days, and was observed before and after diagnosis. Vomiting was more prominent in children. Fecal PCR testing was as accurate as respiratory specimen PCR detection, and fecal excretion persisted after sputum excretion in 23% of the patients for 1 to 11 days.

A systematic review of four case series (36 children, 15 boys and 21 girls, aged 56 to 91 months) aimed to investigate differences in viral shedding in respiratory and fecal samples from children with Covid-19. ^W34^ Three times as many children had SARS-CoV-2 shedding in stools after 14 days of symptoms onset compared to respiratory samples, RR= 3.2 (95%CI 1.2 to 8.9). The review authors were unsure whether the fecally shed viruses were viable and hence infectious.

A short review by Donà et al. ^W14^ including six studies of children reports the orofecal route is an alternative route of transmission, regardless of presenting Covid-19 symptomatology. Citing evidence from the SARS-CoV-1 outbreak that fecal excretion could be ongoing even after 30 days from symptom onset, the review authors recommend that exclusion of SARS-CoV-2 infection by single time point nasopharyngeal swabs should not be used in children.

A review on the potential for foodborne transmission of SARs-CoV-2 found no published studies of SARS-CoV-2 survival in or on food products. ^W20^

## Live SARS-COV-2 excretion by the fecal route

Six studies ([Jeong](https://www.cebm.net/study/viable-sars-cov-2-in-saliva-urine-and-stool-from-covid-19-patients/), ^w18^, [Wang W](https://www.cebm.net/study/detection-of-sars-cov-2-in-different-types-of-clinical-specimens/), ^w41^ [Xiao F Tang M](https://www.cebm.net/study/covid-19-evidence-for-gastrointestinal-infection-by-sars-cov-2-2/), ^w47^ [Xiao F Sun J](https://www.cebm.net/study/infectious-sars-cov-2-in-feces-of-patient-with-severe-covid-19/), ^w48^ [Yoa H](https://www.cebm.net/study/impact-pathogenicity-of-sars-cov-2/), ^w51^ and [Zhang Y](https://www.cebm.net/study/isolation-of-sars-cov-2-from-a-stool-specimen-of-confirmed-case-of-covid-19/) ^w58^) reported isolating the virus from nine patients: one study [Qian](https://www.cebm.net/study/direct-evidence-of-active-sars-cov-2-replication-in-the-intestine/), ^w33^ reported virions in rectal tissue under electron microscopy; and a laboratory study by [Lamers](https://www.cebm.net/study/sars-cov-2-productively-infects-human-gut-enterocytes/) ^w21^ found that SARS-CoV-2 productively infected human small intestinal organoids.

Viable SARS-CoV-2 was isolated from naso/oropharyngeal swabs and saliva of Covid-19 patients, as well as nasal washes of ferrets inoculated with patient urine or stool. ^w18^

Wang W et al ^w41^ detected SARS-CoV-2 in the feces of 44 of 153 (29%) specimens collected from 205 patients with Covid-19 from three hospitals in the Hubei and Shandong provinces and Beijing. RNA was extracted by RT-PCR targeting the open reading frame 1ab gene of SARS-CoV-2. A cycle threshold value less than 40 was interpreted as positive for SARS-CoV-2 RNA. Four positive fecal specimens with high copy numbers were then cultured, and electron microscopy performed to detect live virus. Live SARS-CoV-2 virus was observed in the stool sample of two patients who did not have diarrhea.

Xiao F, Sun J et al. ^w48^ undertook a case-series of 28 hospitalised patients with severe Covid-19. for whom feces samples were available. Among the specimens collected 12 were positive for viral RNA at least one-time point. SARS-CoV-2 virus was successfully isolated from two of the viral RNA–positive patients. Viral particles that were visible were spherical and had distinct surface spike protein projections, consistent with a previously published SARS-CoV2 image. Four serial feces samples from a seriously ill 78-year old patient with Covid-19 all tested positive for viral RNA; the patient subsequently died. Viral antigen was also detected in gastrointestinal epithelial cells of a biopsy sample, from this 78-year old Covid-19 patient.

An assessment of viral RNA in feces from 71 hospitalized patients with SARS-CoV-2 reported that viral RNA and viral nucleocapsid protein in gastrointestinal tissues was extracted from one patient. ^w47^ Immunofluorescent data showed that ACE2 protein (a cell receptor for SARS-CoV-2) is expressed in the glandular cells of gastric, duodenal, and rectal epithelia, supporting the entry into the host cells. Viral nucleocapsid protein was found intracellularly in gastric, duodenal, and rectal epithelia.

Yoa H et al. ^w51^ obtained 11 SARS-CoV-2 viral isolates from patients admitted to Zhejiang University-affiliated hospitals in Hangzhou, China. Three of the viable viral isolates were extracted from stool samples, indicating that the SARS-CoV-2 is capable of replicating in stool samples. At 24 hours post-infection significant decreases in cycle threshold value for all of the viral isolates were observed.

Zhang Y ^w58^ isolated live virus from the stools of one severe pneumonia case, pointing to a possible orofecal spread. There were unclear and sparse clinical details in the study report.

A patient admitted to Zhongnan Hospital of Wuhan University for treatment of a rectal adenocarcinoma had samples taken from enteric sections, mucosa of rectum and ileum. ^w33^ Typical coronavirus virions in rectal tissue were observed under electron microscopy with abundant lymphocytes and macrophages (some SARS-CoV-2 positive) infiltrating the lamina propria. Virions were found in the cytoplasm of intestinal epithelial cells and at electron microscopy. SARS-CoV-2 antigens were confirmed to be expressed on intestinal epithelial cells, lymphocytes and macrophages in the lamina propria.

Zang R et al. ^w54^ undertook a laboratory study that reported that human enterocytes express high ACE2 receptor levels, which could support infection with SARS-CoV-2, and Lamers et al. ^w21^ found that SARS-CoV-2 productively infected Human small intestinal organoids (hSIOs) assessed by qRT-PCR for viral sequences and by live virus titrations on VeroE6 cells.

## Timing of fecal shedding

A retrospective study of 133 hospitalised Covid-19 patients identified 22 whose sputum or fecal samples tested positive, after pharyngeal swabs became negative. [[Chen](https://www.cebm.net/study/sars-cov-2-positive-sputum-and-feces-samples-in-covid-19-patients/) ^w7^] [Cheung](https://www.cebm.net/study/sars-cov-2-infection-in-fecal-samples-from-hong-kong/) et al. ^w10^ reported in a cohort of 59 patients that fecal discharge continues long after respiratory shedding of Covid-19 has ceased. [Gupta](https://www.cebm.net/study/covid-19-persistent-viral-shedding-of-sars-cov-2-in-faeces/) et al. ^w15^ reported that the duration for fecal shedding of viral RNA after clearance of respiratory samples ranged from 1 to 33 days and in one patient was up to 47 days from symptom onset.

In the review by Tian et al. ^W39^ fecal PCR testing was shown to be as accurate as respiratory specimen PCR detection, and fecal excretion persisted after sputum excretion in 23% patients for 1 to 11 days. Donà et al. ^W14^ cites evidence from the SARS-CoV-1 outbreak that fecal excretion could be ongoing even after 30 days from symptom onset, the review authors recommend that exclusion of SARS-CoV-2 infection by single time point nasopharyngeal swabs should not be used in children.

## Detection in bathrooms of Acute Healthcare Settings

From January 24th to February 4th, three infected patients in airborne infection isolation rooms with anterooms and bathrooms had surface environmental samples taken at 26 sites. [Ong](https://www.cebm.net/study/air-surface-environmental-and-personal-protective-equipment-contamination-by-sars-cov-2-from-a-symptomatic-patient/) et al. ^w29^ In two symptomatic patients rooms, after routine cleaning all samples were negative. In the third patient’s room, samples were collected before routine cleaning and were found to be positive for 13 out of 15 (87%) sites (including air outlet fans) and three of five toilet sites (toilet bowl, sink, and door handle) tested positive. One patient had upper respiratory tract involvement and two positive stool samples for SARS-CoV-2 on RT-PCR despite not having diarrhoea. ^w29^

A study in February 2020 randomly sampled the 3-bed isolation rooms of the Covid-19 designated infectious diseases hospital wards, Nanjing, China. [[Ding Z](https://www.cebm.net/study/covid-19-toilets-dominate-environmental-detection-of-sars-cov-2-in-a-hospital/) ^w13^] Environmental sampling was also carried out in four isolation rooms, a nursing station, a corridor, an air-conditioning system and other spaces in the airborne infectious-disease zone on the fifth floor of the hospital. Sampling procedures are described accurately and the air sampler twice needed to be quarantined despite wearing full PPE. Airflow was also assessed between 4th and 5th floors in the building using a smoke tracer. Of the 107 surface samples (37 from toilets, 34 from other surfaces in isolation rooms and 36 from other surfaces outside isolation rooms). Four samples were positive (two ward door door-handles, one bathroom toilet toilet-seat cover and one bathroom door door-handle). Three were weakly positive from a bathroom toilet seat, one bathroom washbasin tap lever and one bathroom ceiling exhaust louvre. One of the 46 corridor air samples was weakly positive.

## Detection in Sewage

[Ahmed](https://www.cebm.net/study/isolation-of-sars-cov-2-in-untreated-wastewater-in-australia/) et al. ^w1^ obtained samples taken from two treatment plants and pumping stations around Brisbane, Queensland, Eastern Australia starting on the 24th of February until 13th of April with more frequent sampling as the epidemic curve got higher. Some 22% of the samples from one sampling point were positive. The estimated number of infections and prevalence were strongly correlated with the log10 SARS-CoV-2 RNA copies in stool, followed by the RNA copies detected in wastewater and log10/g of feces/person/day.

[Cahill](https://www.cebm.net/study/covid-19-in-wastewater/) et al. ^w3^ review of available evidence of the orofecal transmission and its possible relation to wastewater management reported that SARS-CoV-2 has been detected in faeces and wastewater over recent months.

Concentration and detection of SARS coronavirus was found in sewage concentrates in China in two hospitals receiving SARS patients prior to disinfection, and occasionally after disinfection (no live SARS-CoV was detected in these assays). The authors reported that the virus may be able to survive for 14 days in sewage at 4°C, 2 days at 20°C; its RNA could be detected for eight days although the virus had been inactivated. [[Wang XW](https://www.cebm.net/study/concentration-and-detection-of-sars-coronavirus-in-sewage-in-china/) ^w42^]

Sentinel surveillance of SARS-CoV-2 in wastewater was shown to anticipate the occurrence of Covid-19 cases. SARS-CoV-2 was detected in sewage 41 days before the declaration of the first Covid-19 case in Spain and in frozen samples dating back to 12 March 2019. If confirmed, the results hypothesise that SARS CoV-2 has been around much longer than considered. [[Chavarria-Miró G](https://www.cebm.net/study/covid-19-sentinel-surveillance-of-sars-cov-2-in-wastewater-anticipates-the-occurrence-of-covid-19-cases/) ^w6^]

Even at low Covid-19 prevalence, sewage surveillance could be a sensitive tool to monitor the viral circulation. SARS-CoV-2 RNA detection occurred in volumes of 250 mL of wastewaters collected in both areas at high (Milan) and low (Rome) epidemic circulation, according to clinical data. Six out of 12 samples were positive. [[La Rosa](https://www.cebm.net/study/first-detection-of-sars-cov-2-in-untreated-wastewaters-in-italy/) (a) ^w23^] One of the positive results was obtained in a Milan wastewater sample collected three days after the first notified Italian case of autochthonous SARS-CoV-2 at Codogno. A second positive sample was taken on the 28th of February in Milan when Covid-19 cases were only 29.

Environmental screening may, therefore, be a sensitive tool to gauge viral presence before clinical symptoms become apparent. SARS-CoV-2 was detected in the sewage of five sites a week after the first Covid-19 case in the Netherlands: identification of viral antigens occurred when the COVID-19 prevalence was around or even below 1 case per 100,000. Stronger signals were observed when the prevalence was 3.5 cases per 100,000 or more. [[Medema G](https://www.cebm.net/study/sars-coronavirus-2-in-sewage-in-the-netherlands/) ^w27^]

A systematic review of what is known of the presence and survival of coronaviridae in various water settings included twelve articles. [La Rosa](https://www.cebm.net/study/covid-19-coronaviruses-in-water-environments/) et al. ^w22^ noted the methods of concentration and collection for these viridae may not be appropriate. Coroviridae have been isolated in different types of liquids from waste to surface water but in general, they appear to be unstable. Chlorination and higher temperatures lead to their inactivation. Chlorine was considered far more effective against SARS-CoV-2 than other microorganisms because of its lytic action on the envelope.

A study in the First Affiliated Hospital of Zhejiang University, China found that strict disinfection and hand hygiene could decrease the hospital-associated Covid-19 infection risk of the staff in isolation wards. The study monitored the presence of SARS-Cov-2 among hospital environment surfaces, sewage, and personal protective equipment (PPE) of staff. During the study period, 33 patients were hospitalized in isolation wards, and no SARS-Cov-2 RNA was detected among the 36 objects surface samples and nine staff PPE samples. [Wang J (b)](https://www.cebm.net/study/covid-19-hygiene-monitoring-during-the-coronavirus-disease-2019-outbreak-in-a-chinese-hospital/) et al. ^w41^ surfaces were wiped with 1000 mg/L chlorine-containing disinfectant every 4 hours in the isolation ICU ward and every 8 hours in general isolation wards. Preprocessing disinfection equipment was added before sewage drainage from the isolation wards into the final sewage disinfection pool. The sewage samples from the inlet pool were found to be positive, and from the outlet of the preprocessing pool weakly positive, but the sewage samples from the last disinfection pool were negative. No viable virus was detected by culture.

## Discussion

We found nine reviews and 51 primary studies or reports examining the potential role of orofecal transmission of SARS-CoV-2. Thirty seven studies reported positive fecal samples for SARS-CoV-2 based on RT-PCR results of 1,034 patients. Six studies reported isolating the virus from fecal samples of nine patients and one study from rectal tissue. Eleven studies report on fecal samples found in sewage, and two collected samples from bathrooms and toilets.

## How this fits with knowledge of other Coronaviruses

MERS-CoV has been shown to infect human primary intestinal epithelial cells, small intestine explants and intestinal organoids.^8^ MERS-CoV has been detected in 42% of milk samples collected from lactating camels where it can survive for a prolonged period. A study of human primary intestinal epithelial cells and small intestine explants of MERS-CoV patterns identified the viral replication intermediates in stool specimens. MERS-CoV was found to be resistant to fed-state gastrointestinal fluids but less tolerant to the high acidic fasted-state gastric fluid.

In 1977 the prolonged excretion of coronaviruses in feces was first observed.^10^ In the SARS-CoV-1 outbreak in 2002-03, a significant portion of patients had enteric involvement. In the Toronto outbreak in 2003, 6% of 144 patients had diarrhoea on presentation.^11^ Also among 138 patients with SARS in Hong Kong, 20% presented with watery diarrhoea and 38% had symptoms of diarrhoea during the illness. Intestinal biopsy specimens showed the presence of active viral replication, and SARS-CoV RNA was detected in the stool of some patients for more than ten weeks after symptom onset.^12^ A retrospective study on specimens from 154 patients in Hong Kong with laboratory-confirmed SARS found the viral load to be the highest in stool specimens.^13^ Up to 70% of 75 patients in a community outbreak in Hong Kong developed watery diarrhea.^14^ This outbreak was linked to a faulty sewage system in the Amoy Gardens apartment complex, further suggesting orofecal transmission might be a route for transmission.^15^

The human gastrointestinal tract can act as a primary infection site for SARS-CoV. Ding et al used a monoclonal antibody specific for the SARS-CoV nucleoprotein, and probes for the RNA polymerase gene fragment in four patients who died from SARS-CoV-1.^16^ Virus was detected in the stomach, small intestine, distal convoluted renal tubule, sweat gland, parathyroid, pituitary, pancreas, adrenal, liver and cerebrum. The authors discussed that viruses in contaminated food and water may enter the human body through epithelial cells covering the surface of the gastrointestinal tract, although there was no direct evidence to show that food-borne transmission had occurred. A study from the sewage of two hospitals receiving SARS patients in Beijing found no infectious SARS-CoV contamination in any of the samples collected, but did detect the nucleic acid in the sewage from the two hospitals before disinfection - providing further evidence that SARS-CoV-1 can be excreted by feces into the sewage system. ^17^

While human coronaviruses are considered not to transmit fecally this is not the case in animals. Feline coronavirus, for instance, is typically shed in feces of healthy cats and transmitted by the orofecal route to other cats.^19^ Pigs are also infected by the transmissible gastroenteritis coronavirus via the fecal-oral route.^20^ Bat coronavirus infects the gastrointestinal and respiratory tracts of bats seemingly without causing disease.^21^ Transmission following exposure to camel feces has also been considered to be biologically plausible, although no evidence indicates whether this is possible. ^22^

There is, however, evidence that SARS-CoV-2 can survive adverse conditions in the gastrointestinal system. It has been identified in endoscopic specimens of the esophagus, stomach, duodenum, and rectum of Covid-19 patients; substantial amounts of SARS-CoV-2 RNA have been consistently detected in stool specimens. The Ding S et al. ^w12^ review cited evidence that SARS-CoV-2 can survive the adverse conditions in the gastrointestinal system. Heavy glycosylation of the large spike S protein has been shown to lead to resistance to the proteases, the low pH and bile salts found in the gastrointestinal system. Some gastric processes may actually facilitate viral entry into the enterocytes: in bovine coronavirus, one specific site on the S glycoprotein has to be cleaved by an intracellular protease or trypsin to activate viral infectivity and cell fusion.^3^

## Limitations

The limitations in this review are mainly related to the quality of the included studies. Many were small, did not provide a protocol that established a priori methods, they were often poorly reported and often did not take into account biases. These limitations prevent firm conclusions being drawn on the magnitude of the estimates but these studies do provide largely consistent evidence on the main conclusions that SARS-CoV-2 is excreted fecally, is found in sewage and can be cultured.

## Implications for Policy

Policy should emphasise routine surveillance of food, wastewaters and effluent. The importance of strict personal hygiene measures, chlorine-based disinfection of surfaces in locations with presumed or known SARS CoV-2 activity should form part of public policy and education campaigns. Stool testing should be carried out in dischargees from the hospital or other holding facilities well before discharge date and discharge should be conditional either on cessation of fecal excretion or strict quarantine and personal hygiene measures in those still excreting viral particles by stool independently from respiratory excretion.

## Implications for Further Research

Each outbreak should be investigated and a report be made publicly available rapidly. Testing of stools should be carried out in all people involved in the outbreak. As there is coherent evidence of ingestion, penetration of enterocytes and excretion of live SARs-CoV-2 in possible analogy with SARs and MERS agents, we believe this working hypothesis should be tested by conducting case-control studies during the investigation of outbreaks following a set protocol. For such investigations, cases would be cases of Covid-19 (categorised by symptom presence and severity) either fecally excreting virions or not (cases and contacts) and controls would be healthy matches. Exposure to potentially fecally contaminated materials and protective measures taken would be elicited at interview. To minimise the play of recall and ascertainment bias, interviewers should be blind to fecal excretion status and the interview should take place as soon as possible after the event.

Viability of fecal isolates and their possible pathogenicity should be tested in outbreaks, irrespective of the presence of symptoms or nasal swab positivity.

## Conclusion

The various observational and mechanistic evidence supports the hypothesis that SARS-CoV-2 can infect and be shed from the human gastrointestinal tract. This has important implications for policy that should emphasise routine surveillance of food, wastewaters and effluent, and emphasize the importance of strict personal hygiene measures, chlorine-based disinfection of surfaces in locations with presumed or known SARS CoV-2 activity.

## Data Availability

Thje data is avialbe in our open evidence review: Analysis of the Transmission Dynamics of COVID-19: An Open Evidence Review, that is investigating factors and circumstances that impact on the transmission of SARS-CoV-2, based on our published protocol.

https://www.cebm.net/evidence-synthesis/transmission-dynamics-of-covid-19/

## Funding

This review received funding support from the National Institute of Health Research School of Primary Care Research (NIHR SPCR) Evidence Synthesis Working Group Project 380. and Funding from the David and Maria Willetts Foundation.

## Acknowledgment

Drs Susan Amirian, Siyuan Ding and Sravanthi Parasato provided additional information for this review.

## Authors

Tom Jefferson is a senior associate tutor and honorary research fellow, Centre for Evidence-Based Medicine, University of Oxford. Disclosure statement is [here](https://restoringtrials.org/competing-interests-tom-jefferson/)

Elizabeth Spencer is Epidemiology and Evidence Synthesis Researcher at the Centre for Evidence-Based Medicine. ([Bio here](https://www.phc.ox.ac.uk/team/elizabeth-spencer))

Jon Brassey is the Director of Trip Database Ltd, Lead for Knowledge Mobilisation at Public Health Wales (NHS) and an Associate Editor at the BMJ Evidence-Based Medicine.

Carl Heneghan is Professor of Evidence-Based Medicine, Director of the Centre for Evidence-Based Medicine and Director of Studies for the Evidence-Based Health Care Programme. (Full bio and disclosure statement [here](https://www.phc.ox.ac.uk/team/carl-heneghan))

## Disclaimer

The article has not been peer-reviewed. The views expressed in this commentary represent the views of the authors and not necessarily those of the host institution, the NHS, the NIHR, or the Department of Health and Social Care. The views are not a substitute for professional medical advice. It will be regularly updated see the evidence explorer at https://www.cebm.net/evidence-synthesis/transmission-dynamics-of-covid-19/ for regular updates to the evidence summaries and briefs.

This work is licensed under a [Creative Commons Attribution-NonCommercial 4.0 International License](http://creativecommons.org/licenses/by-nc/4.0/).

## Notes

### Competing Interest Statement

The authors have declared no competing interest.

### Author Declarations

no IRB/oversight required.

